# In children with transfusion-dependent thalassemia, inflammation and neuronal damage biomarkers are associated with affective and chronic fatigue symptoms

**DOI:** 10.1101/2023.11.21.23298798

**Authors:** Maha Abdul Saheb Ridhaa, Hussein Kadhem Al-Hakeim, Mohammed K. Kahlol, Tabarek Hadi Al-Naqeeb, Michael Maes

**Author notes:** Correspondence Author: Prof.Dr. Michael Maes, Sichuan Provincial Center for Mental Health, Sichuan Provincial People’s Hospital, School of Medicine, University of Electronic Science and Technology of China, Chengdu 610072, China. https://scholar.google.co.th/citations?user=1wzMZ7UAAAAJ&hl=th&oi=ao.

## Abstract

**Background:** Patients with transfusion-dependent thalassemia (TDT) are vulnerable to neurotoxicity due to frequent blood transfusions and the subsequent iron overload (IO) and inflammation. As a result, affective (depression and anxiety) and chronic fatigue syndrome (CFS) symptoms may develop.

**Aims:** To investigate the potential association between TDT and neuronal injury, as assessed with serum concentrations of neuronal damage biomarkers, including neurofilament light (NFL), glial fibrillary acidic protein (GFAP), neuron-specific enolase (NSE), and nestin.

**Methods:** We investigated the associations between those CNS injury biomarkers, neuro-immune markers (C-reactive protein (CRP), interleukin (IL)-6, and IL-10), calcium, magnesium, copper and zinc, and the Fibro-Fatigue (FF), the Children’s Depression Inventory (CDI), and the Spence Children’s Anxiety Scale (SCAS) scores in 126 children with TDT and 41 healthy children.

**Results:** TDT children show significant increases in IO, FF, CDI, and SCAS scores, serum NSE, GFAP, NF-L, CRP, copper, IL-6, and IL-10, and lowered magnesium, zinc, and calcium as compared with healthy children. There were significant correlations between the CDI score and NFL, NSE and GFAP; SCAS score and NFL, and FF score and NFL and GFAP. The neuronal damage biomarkers (except nestin) were significantly associated with inflammatory, erythron (hematocrit and hemoglobin) and IO (iron and ferritin) biomarkers.

**Conclusions:** TDT is characterized by intertwined increases in neuronal injury biomarkers and neuropsychiatric symptoms suggesting that TDT-associated neurotoxicity plays a role in affective symptoms and CFS due to TDT. Inflammation and neurotoxicity are novel drug targets for the prevention of affective symptoms and CFS due to TDT.

## Introduction

Beta-thalassemia (β-TM) encompasses a group of hereditary conditions characterized by the absence or deficiency of the β-globin chain in the hemoglobin A molecule. This chain defect causes harm to the erythrocyte membrane and ultimately leads to anemia (Raguram et al., 2022). The aforementioned condition often presents with symptoms of hemolytic anemia and has a 15% mortality rate among the millions of children it impacts (Raguram et al., 2022, Crippa et al., 2019). Individuals with β-TM necessitate long-term blood transfusions to maintain adequate hemoglobin levels and mitigate the adverse consequences of erythropoiesis inefficiency (Aydinok, 2020). Transfusion-dependent thalassemia (TDT) patients are at risk of developing various complications because of requiring frequent blood transfusions (Shah et al., 2019). Prolonged blood transfusions have the potential to cause substantial iron overload (IO), which can result in toxicity affecting various organs such as the heart, liver, and endocrine glands (Hamed et al., 2016, Daher et al., 2017). Iron in excess has been found to be neurotoxic due to its capacity to trigger oxidative stress within the brain (Schipper, 2004). Neuronal death and neurodegeneration (Schneider and Bhatia, 2013), neuroinflammation, Parkinson’s disease and Alzheimer’s disease (Stankiewicz et al., 2007, Hagemeier et al., 2012), neuropsychiatric disorders including affective symptoms and chronic fatigue syndrome (CFS) (Beard et al., 2005, Maaroufi et al., 2009, Al-Hakeim et al., 2020, Al-Hakeim et al., 2021), and cognitive impairments (Wang, 2013) are all potential consequences of IO in the brain. Neuroprotection may result from the use of iron chelators to decrease excessive iron deposition (Ward et al., 2014).

Thalassemia is associated with a heightened prevalence of neuropsychiatric disorders (Sahu et al., 2023). Around sixty-five percent (Sahu et al., 2023) to eighty-three percent (Shaligram et al., 2007) of children with thalassemia may suffer from anxiety disorders, depression, or oppositional defiant disorder. Anxiety symptoms were present in 32% of thalassemia patients and major depression (MDD) in 16-31% (Gan et al., 2016, Shafiee et al., 2014). MDD is associated with elevated IO and pro-inflammatory cytokine levels in the serum, such as interleukin (IL)-1β and tumor necrosis factor (TNF)-α, which are indicators of immune activation in TDT children (Al-Hakeim et al., 2020). Blood transfusions may induce depressive symptoms and MDD in TDT via the mediating effects of IO and subsequent activation of the immune-inflammatory response system (IRS), according to the conclusion reached by the latter authors. MDD has been linked to alterations in iron metabolism, as indicated by elevated serum ferritin levels, decreased erythrocyte count, and decreased hematocrit and hemoglobin concentrations (Rybka et al., 2013, Trevisan et al., 2016).

Additionally, evidence suggests that the impacts of activated IRS pathways on MDD (Schiepers et al., 2005, Maes, 2008, Maes et al., 2022b, Al-Hakeim et al., 2020) and CFS are mediated by heightened neurotoxicity (Maes et al., 2022a). MDD is distinguished by increased concentrations of neurofilament light chain (NFL), glial fibrillary acidic protein (GFAP), P-tau217, and IRS biomarkers including C-reactive protein (CRP) (Al-Hakeim et al., 2023a). NFL levels are elevated not only in patients with MDD and bipolar disorder (Al-Hakeim et al., 2020, Chen et al., 2022) but also in those with treatment-resistant depression (Domingues et al., 2019, Spanier et al., 2019). Serum concentrations of NFL, nestin, CRP, and IL-10 are increased in patients with end-stage renal disease, while serum levels of zinc levels are reduced in comparison to the control group (Al-Hakeim et al., 2023b). NFL is a biomarker of neuronal injury and neurodegenerative processes in the central nervous system (CNS), whilst neurodegenerative diseases are frequently associated with elevated NFL plasma levels (Maes et al., 2022c, Yuan and Nixon, 2016). GFAP serves as an indicator of astrocyte functionality, and both its upregulation and rearrangement are crucial factors in the pathogenesis and progression of neurodegeneration, ischemic stroke, and various other CNS disorders (Bagheri et al., 2022, Hol and Pekny, 2015, Yang and Wang, 2015). Two other neuronal injury biomarkers that were not explored in TDT are nestin and neuron-specific enolase (NSE). Nestin mediates radial axonal growth and is an additional intermediate filament protein that is expressed by neuronal progenitor cells, including those in the adult brain (Hendrickson et al., 2011, Paquet-Durand and Bicker, 2007). NSE is an additional neuronal marker that has been linked to neurodegenerative, inflammatory, hypoxic, and metabolic diseases (Park et al., 2019, Bagheri et al., 2022, Haque et al., 2018).

Additionally, magnesium deficiency may contribute to neurodegenerative processes (Hlásný, 2020), and dysfunctional calcium signaling may be one of the essential processes in neurodegenerative processes (Bezprozvanny, 2010). This is significant because IRS activation is associated with decreased calcium, magnesium, and zinc levels and increased copper levels in MDD (Al-Dujaili et al., 2019, Al-Hakeim et al., 2022). Neurodegenerative pathways, including aberrations in the Wnt/catenin pathway, oxidative stress pathways, and IRS activation, may be implicated in CFS (Maes et al., 2022a).

Therefore, we postulated that IO could potentially induce heightened neurotoxicity and, consequently, increased levels of neuronal and astroglial projection lesions in TDT, thereby exacerbating the severity of affective symptoms and CFS. Therefore, the primary objective of the present investigation is to determine whether TDT is correlated with neuronal and astroglial injury indicators (as measured by serum GFAP, NSE, NFL, and nestin), as well as whether these indicators are associated with IRS biomarkers (as measured by CRP, IL-6, and IL-10), affective symptoms, and CFS.

### Subjects and Methods

#### Participants

A total of 167 children were enrolled in the present study; of these, 126 were children with TDT and forty-one were healthy control children between the ages of 6 and 12, of both sexes. The recruitment process for TDT patients commenced in March 2023 and concluded in May 2023 at the Thalassemia Unit, which is situated at Al-Zahra’a Teaching Hospital in the Najaf Governorate of Iraq. The diagnosis of β-TM was ascertained through the utilization of the diagnostic criteria specified in the 2019 ICD-10-CM Diagnosis Code D56.1 by pediatricians and hematologists. The identification of β-TM was accomplished through the assessment of distinctive clinical manifestations, including aberrant bone growth, hepatosplenomegaly, and severe anemia. Hematological assessments were performed, including the measurement of hemoglobin (Hb) concentrations below 70 g/L and the identification of a high proportion of reticulocytes and hypochromic microcytic red blood cells with anisopoikilocytosis. Furthermore, the concentration of HbA2 was ascertained via high-performance liquid chromatography (HPLC) employing the VARIANT TM β-Thalassemia Short Program.

The control group comprised forty-one infants who were in good health. There were no cases of anemia, immune-inflammatory disorders, or systemic diseases among the controls. Children with splenectomy, systemic diseases including renal failure, or diabetes mellitus were precluded from the study. The determination of when to administer packaged red blood cells (RBCs) for blood transfusions was based on the Hb levels, which had to be maintained at or above 90 g/L. This was accomplished every two to four weeks. Furthermore, a protocol of iron-chelating therapy was implemented for the patients. This involved administering deferoxamine mesylate USP (brand name Desferal^®^) infusions at a rate ranging from 25 to 50 mg/kg/day for a duration of eight hours daily. The precise dosage that was administered to the patients was ascertained by analyzing their blood ferritin levels. To alleviate the symptoms of ineffective erythropoiesis, folic acid was administered to most patients undergoing TDT. Vitamin C was administered to patients who had received a diagnosis of TDT as a therapeutic intervention to support the iron chelation process when used in conjunction with deferoxamine.

The researchers obtained written informed consent from the first-degree relatives (either the father or mother) of the participating children after a comprehensive verbal explanation, in adherence to the tenets delineated in the Declaration of Helsinki. The research was granted approval by the Institutional Review Board (IRB) of the Najaf Health Directorate, Training and Human Development Center, Iraq (document number 4233/2023).

#### Clinical measurements

The Fibro-fatigue (FF) scale was administered by a senior psychiatrist to evaluate the severity of CFS in both patients and controls (Zachrisson et al., 2002). The assessment of depressive symptoms was conducted utilizing the Children’s Depression Inventory (CDI). This is a self-rating screening instrument consisting of twenty-seven three-point scale statements. The child is requested to choose the response that most accurately characterizes his or her emotions over the past two weeks in relation to each given statement. The total CDI score is utilized as an indicator of depression severity in the present study. Negative self-esteem, interpersonal difficulties, ineffectiveness, anhedonia, and negative mood are the subscales that comprise the CDI. Having been validated in school-aged children, the CDI is one of the most widely utilized depression screening instruments for children. Depression in children may be diagnosed when the Child Depression Inventory (CDI) score is equal to or greater than nineteen. The children’s anxiety symptoms were assessed using the Spence Children’s Anxiety Scale (SCAS) (Spence et al., 2003).

#### Assays

Five milliliters of venous blood were drawn from all participants after an overnight fast. The patients’ samples were collected just before their blood transfusion session. Blood was left until complete clotting (about ten minutes) at room temperature. After centrifugation for five minutes at 3000 rpm, serum was separated and transported into Eppendorf tubes. Serum calcium, magnesium, albumin, copper, and zinc were measured by using ready-for-use kits purchased from Agappe Diagnostics^®^ ready-to-use kit from Cham (Switzerland). Copper was determined spectrophotometrically using a kit from LTA Co. (Milano, Italy). The amount of iron in sera was determined by colorimetric kits purchased from Spectrum^®^ (Cairo, Egypt). Commercial ELISA sandwich kits provided by Nanjing Pars Biochem Co., Ltd., Nanjing, China, were used to measure interleukin-6, Interleukin-10, GFAP, NSE, NF-L, and nestin. Serum ferritin levels were measured by using an ELISA kit purchased from Elabscience^®^ (Wuhan, China). These kits were made for human samples depending on the biotin double antibody sandwich technology. Hematological parameters were measured by a five-part differential Mindray BC-5000 hematology analyzer (Mindray Medical Electronics Co., Shenzhen, China). The inter-assay CV% of all kits is lower than 10%. For samples with highly concentrated analytes, we employed sample dilutions. A C-reactive protein (CRP) latex slide test (Spinreact^®^, Barcelona, Spain) was used for CRP measurement in human serum. The approximate CRP concentration in the patient sample is calculated as follows: 6 x CRP Titer = mg/L.

#### Statistical analysis

Analysis of contingency tables (χ^2^ test) was employed to examine the relationships between nominal variables, whereas analysis of variance (ANOVA) was utilized to assess variations in continuous variables across groups. To determine the correlations between scale variables, Pearson’s product-moment correlation coefficients were applied. General linear models (GLM) were employed in the current investigation to assess the correlations between TDT (in contrast to the control group) and the biomarkers. Following that, evaluations of between-subject relationships and pairwise comparisons were performed across treatment groups. The researchers utilized a false-discovery rate (FDR) procedure to mitigate the impact of type I errors that may occur during multiple comparisons (Benjamini and Hochberg, 1995). Multiple regression analysis was utilized in the study, employing both manual and automatic stepwise techniques. A p-to-entry threshold of 0.05 and a p-to-remove threshold of 0.06 were applied. The purpose of the analysis was to determine which biomarkers could be used to predict the scores by assessing the variation in R^2^ (used as effect size). To identify (multi)collinearity issues, tolerance, and VIF were employed, Cook’s distance and leverage were utilized to assess multivariate normality, and the White and modified Breusch-Pagan tests were utilized to determine homoscedasticity. The results of the regression analyses were bootstrapped using 5,000 samples, and when there was an incongruity between the outcomes, the bootstrapped results were displayed. Four z-unit-based composite ratings were computed. a) IRS activation was calculated as z IL-6 + z IL-10 + z CRP - z zinc – z albumin (referred to as Comp_IRS); b) Disorders in the erythron were represented by z hemoglobin + z hematocrit (labeled as Comp_erythron); c) z Iron + z ferritin constituted Comp_IO; and d) an overall severity index was determined as z FF + z CDI + z SCAS (labeled as Comp_severity). We extracted the first principal component (PC) using the principal component (GFAP), NSE, and Nestin (NFL could not be included because it did not load significantly on the first PC). The first principal component (PC) accounted for 50.3% of the variance, and all loadings exceeded 0.67. These results suggest that the PC score, designated PC_neuronal, potentially reflects neuronal injury. All statistical tests were two-tailed, and a p-value of 0.05 was used for statistical significance. For the analysis of the data, version 29 of IBM SPSS for Windows was utilized.

## Results

### 1. Socio-demographic data

**Table 1** shows the socio-demographic and clinical data of the children participating in the current study. The TDT patients have a duration of disease (appearance of severe symptoms) of 8.84±2.75 years. There were no significant differences in age, sex ratio, BMI, and education years between healthy children and TDT patients. The results showed significant increases (all p<0.001) in total FF, CDI, and SCAS scores, ferritin, and iron in TDT patients compared with healthy children. There was a significant decrease in Hb and PCV% in patients compared with the controls.

**Table 1.**
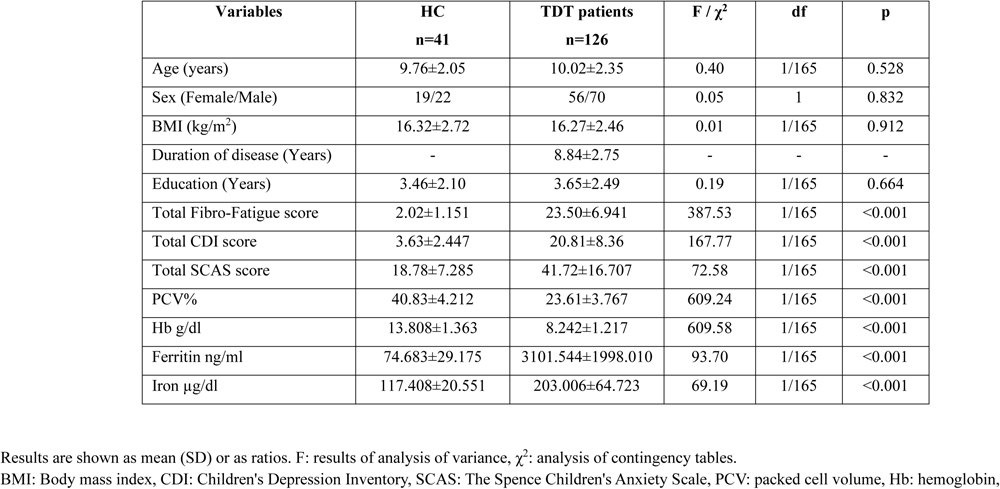
Baseline characteristics of transfusion-dependent thalassemia (TDT) patients in comparison with the healthy controls (HC)

### 2. Biomarkers in TDT and controls

**Table 2** shows the results of the biomarkers in TDT and control groups. The results showed a significant increase in the serum CRP, copper, IL-6, IL-10, NSE, GFAP, and NFL in the TDT group compared with the healthy children. The Comp_IRS and PC_neuronal scores were both significantly increased in TDT patients compared with the control group. Magnesium, calcium, and zinc are significantly decreased in TDT compared with the control group. These differences remained significant after FDR’s p-correction. Serum albumin and nestin are not significantly different between the diagnostic groups.

**Table 2.** Serum biomarkers in transfusion-dependent thalassemia (TDT) patients in comparison with the healthy controls (HC)

| Variables | HC<br>n=41 | TDT patients<br>n=126 | F | df | p |
| --- | --- | --- | --- | --- | --- |
| C-reactive protein mg/l | 4.00±2.03 | 16.13±13.31 | 67.14 | 1/165 | <0.001 |
| Albumin g/dl | 4.368±0.699 | 4.274±0.697 | 0.56 | 1/165 | 0.455 |
| Magnesium mM | 1.269±0.295 | 1.058±0.253 | 19.65 | 1/165 | <0.001 |
| Calcium mM | 2.484±0.218 | 2.218±0.221 | 45.16 | 1/165 | <0.001 |
| Copper mg/l | 0.974±0.162 | 1.329±0.500 | 19.95 | 1/165 | <0.001 |
| Zinc mg/l | 0.761±0.184 | 0.641±0.192 | 13.54 | 1/165 | <0.001 |
| IL-6 pg/ml | 4.768±3.483 | 7.554±4.768 | 11.92 | 1/165 | 0.001 |
| IL-10 pg/ml | 7.083±2.929 | 16.154±11.582 | 34.33 | 1/165 | <0.001 |
| Neuron-specific enolase ng/ml | 0.787±0.563 | 1.291±1.442 | 7.33 | 1/165 | 0.008 |
| GFAP pg/ml | 2.972±2.348 | 4.451±4.789 | 5.85 | 1/165 | 0.017 |
| NFL ng/ml | 1.307±1.243 | 2.980±2.246 | 32.59 | 1/165 | <0.001 |
| Nestin ng/ml | 34.126±17.051 | 42.167±30.873 | 1.44 | 1/165 | 0.231 |
| Comp_IRS (z score) | -0.983±0.629 | 0.320±0.884 | 76.40 | 1/165 | <0.001 |
| PC_neuronal (z score) | -0.399±0.696 | 0.130±1.051 | 9.08 | 1/165 | 0.003 |

wn as mean (SD) or as ratios. F: results of analysis of variance; GFAP: Glial fibrillary acidic protein, NFL: neurofilament light chain, IL: PC: principal components; Comp: composite score, IRS: immune-inflammatory responses system.

### 3. Correlation matrix between clinical scores and biomarkers in TDT

**Table 3** shows the correlation matrix of the neuropsychiatric rating scale scores and the iron status biomarkers in TDT patients. The results showed that FF is significantly correlated with CRP, copper, IL-6, IL-10, GFAP, and NFL, whilst there is a significant inverse correlation between FF and magnesium, calcium, and zinc. The results indicated that the CDI score is significantly correlated with CRP, IL-10, NSE, GFAP, and NFL, and inversely with magnesium, calcium, and zinc. The SCAS score is significantly correlated with CRP, copper, IL-10, and NFL, and inversely with calcium and zinc. The Comp_erythron index was significantly and inversely correlated with CRP, copper, IL-6, IL-10, NSE, GFAP, and NFL, and positively with magnesium, calcium, and zinc. Comp_IO is significantly correlated with CRP, copper, IL-6, IL-10, NSE, GFAP, and NFL, and inversely with magnesium and calcium.

**Table 3.** Correlation matrix between clinical rating scale scores and biomarkers in transfusion-dependent thalassemia patients and controls.

|  | FF | CDI | SCAS | Comp_erythron | Comp_IO |
| --- | --- | --- | --- | --- | --- |
| C-Reactive protein | 0.496<sup>***</sup> | 0.446<sup>**</sup> | 0.295<sup>***</sup> | -0.490<sup>***</sup> | 0.536<sup>***</sup> |
| Albumin | -0.045 | -0.075 | -0.04 | 0.022 | 0.084 |
| Magnesium | -0.307<sup>***</sup> | -0.262<sup>**</sup> | -0.12 | 0.319<sup>***</sup> | -0.225<sup>**</sup> |
| Calcium | -0.355<sup>***</sup> | -0.335<sup>**</sup> | -0.199<sup>**</sup> | 0.411<sup>***</sup> | -0.385<sup>***</sup> |
| Copper | 0.424<sup>***</sup> | 0.332<sup>**</sup> | 0.315<sup>***</sup> | -0.323<sup>***</sup> | 0.497<sup>***</sup> |
| Zinc | -0.174<sup>*</sup> | -0.204<sup>**</sup> | -0.220<sup>**</sup> | 0.231<sup>**</sup> | -0.078 |
| Interleukin-6 | 0.194<sup>*</sup> | 0.095 | 0.056 | -0.225<sup>**</sup> | 0.237<sup>**</sup> |
| Interleukin-10 | 0.346<sup>***</sup> | 0.361<sup>***</sup> | 0.159<sup>*</sup> | -0.389<sup>***</sup> | 0.375<sup>***</sup> |
| Neuron-specific enolase | 0.096 | 0.163<sup>*</sup> | -0.083 | -0.221<sup>**</sup> | 0.172<sup>*</sup> |
| Glial fibrillary acidic protein | 0.203<sup>**</sup> | 0.198<sup>**</sup> | -0.018 | -0.192<sup>*</sup> | 0.266<sup>***</sup> |
| Neurofilament light | 0.322<sup>***</sup> | 0.288<sup>***</sup> | 0.333<sup>***</sup> | -0.361<sup>***</sup> | 0.366<sup>***</sup> |
| Nestin | -0.002 | -0.003 | 0.022 | -0.055 | 0.144 |

.01, \*\*\*p<0.001 : composite based on hematocrit and hemoglobin, Comp\_IO: composite based on iron and ferritin reflecting iron overload, FF: Fibro-Fatigue score, Depression Inventory score, SCAS: The Spence Children's Anxiety Scale score.

There were significant correlations between Comp_severity and Comp_IRS (r=0.349, p<0.001), Comp_IO (r=0.275, p<0.001) and Comp_erythron (r=-0.221, p<0.001).

### 4. Prediction of neuropsychiatric scores

**Table 4** shows the results of multiple regression analyses with the total FF, CDI, and SCAS scores as dependent variables and the neuronal damage biomarkers and other biomarkers as explanatory variables. The first regression shows that 42.9% of the variance in the FF score was predicted by CRP, copper, and NFL (all positively), and calcium and magnesium (both inversely associated). Regression #2 shows that the CDI total score was predicted by CRP, IL-10, and NFL (all three positively) and inversely with calcium. The third regression shows that 23.2% of the variance in the SCAS total score was explained by NFL, CRP, and copper (all positively correlated), and zinc (inversely associated). **Figure 1** shows the partial regression of the total SCAS scale score on NFL levels.

**Figure 1.**
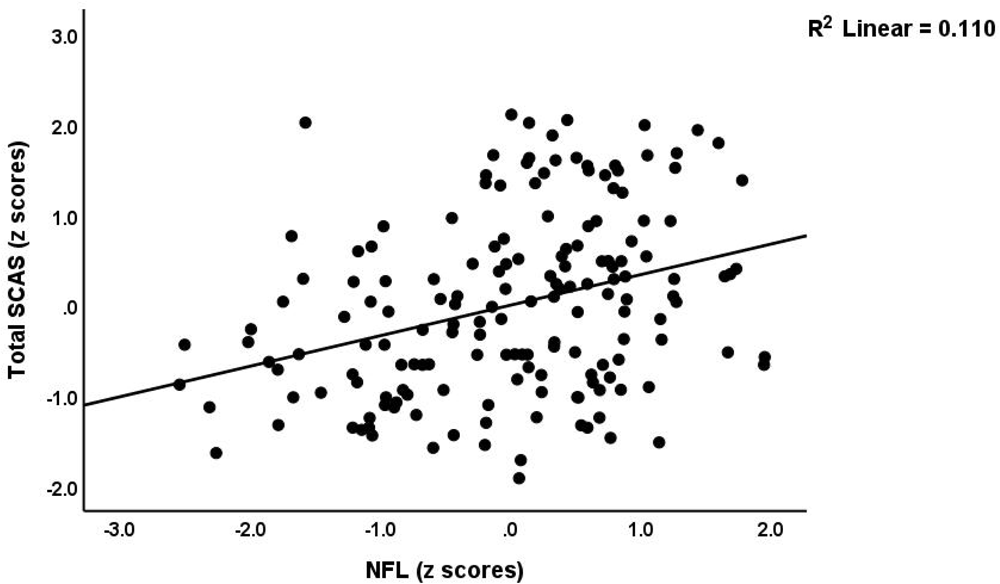
Partial regression of the total Spence Children’s Anxiety Scale (SCAS) on neurofilament light (NFL) levels, after adjusting for age and sex (p<0.001)

**Table 4.** Results of multiple regression analysis with clinical rating scale scores as dependent variables and biomarkers as explanatory variables.

| Dependent Variables | Explanatory Variables | $\beta$ | t | p | F model | df | p | R<sup>2</sup> |
| --- | --- | --- | --- | --- | --- | --- | --- | --- |
| **#1 Fibro-Fatigue score** | **Model #1** |  |  |  | 24.21 | 5/161 | <0.001 | 0.429 |
|  | C-Reactive protein | 0.307 | 4.70 | <0.001 |  |  |  |  |
|  | Copper | 0.270 | 4.16 | <0.001 |  |  |  |  |
|  | Magnesium | -0.187 | -2.99 | 0.003 |  |  |  |  |
|  | Calcium | -0.169 | -2.68 | 0.008 |  |  |  |  |
|  | Neurofilament Light | 0.140 | 2.21 | 0.029 |  |  |  |  |
| **#2. CDI-total score** | **Model #2** |  |  |  | 18.89 | 4/162 | <0.001 | 0.318 |
|  | C-Reactive protein | 0.308 | 4.41 | 0.000 |  |  |  |  |
|  | Interleukin-10 | 0.193 | 2.77 | 0.006 |  |  |  |  |
|  | Calcium | -0.197 | -2.90 | 0.004 |  |  |  |  |
|  | Neurofilament Light | 0.155 | 2.29 | 0.024 |  |  |  |  |
| **#3. SCAS total score** | **Model #3** |  |  |  | 12.24 | 4/162 | <0.001 | 0.232 |
|  | Neurofilament Light chain | 0.214 | 2.93 | 0.004 |  |  |  |  |
|  | C-Reactive Protein | 0.164 | 2.25 | 0.026 |  |  |  |  |
|  | Copper | 0.223 | 3.00 | 0.003 |  |  |  |  |
|  | Zinc | -0.191 | -2.71 | 0.007 |  |  |  |  |

Depression Inventory, SCAS: The Spence Children's Anxiety Scale

### 5. Prediction of the neuronal damage biomarkers by the measured biomarkers in TDT patients

The results in **Table 5** reveal that neuronal damage biomarkers may be predicted by IRS biomarkers. Regression #1 shows that 9.5% of the variance in the NSE could be explained by IL-10. In Regression #2, a part of the variance (12.6 7%) in GFAP is explained by IL-6 and IL-10. **Figure 2** shows the partial regression of GFAP on Comp_IRS. The third regression shows that 16.5% of the variance in NFL is explained by copper (positively) and hemoglobin (inversely). **Figure 3** shows the regression of the NFL levels on hemoglobin. Regression #4 shows that 10.6% of the variance in nestin is positively associated with IL-6 and inversely with calcium. In Regression #5, a moderate part of the variance (24.2%) in PC_neuronal was explained by IL-6, IL-10, and ferritin. Regression #6 shows that 27.1% of the variance in Comp_IRS could be explained by ferritin and inversely by PCV%. **Figure 4** shows the partial regression of Comp_IRS on Comp_erythron.

**Table 5.**
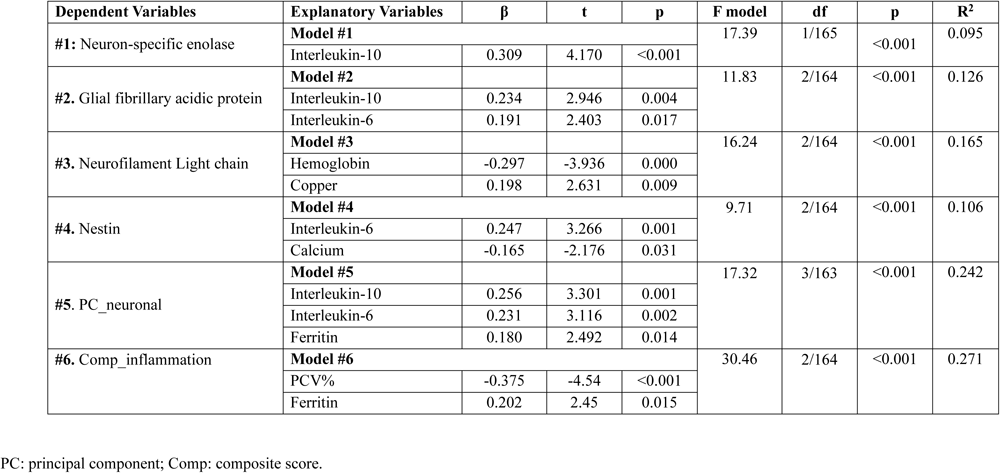
Prediction of the neuronal biomarkers by biomarkers in transfusion-dependent thalassemia patients.

**Figure 2.**
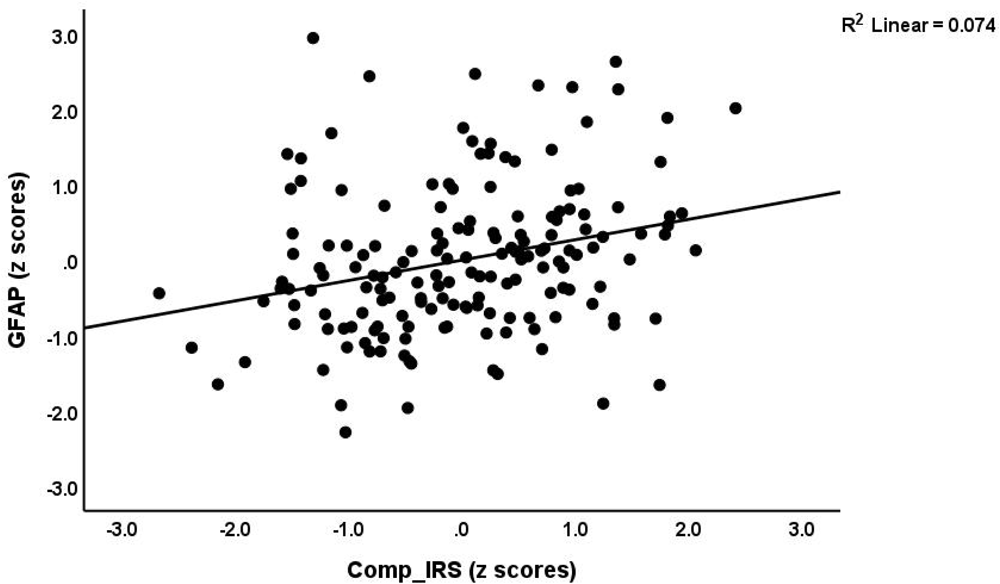
Partial regression of the glial fibrillary acidic protein (GFAP) on an index of immune-inflammatory response system (Comp_IRS) activation, after adjusting for age and sex (p< 0.001)

**Figure 3.**
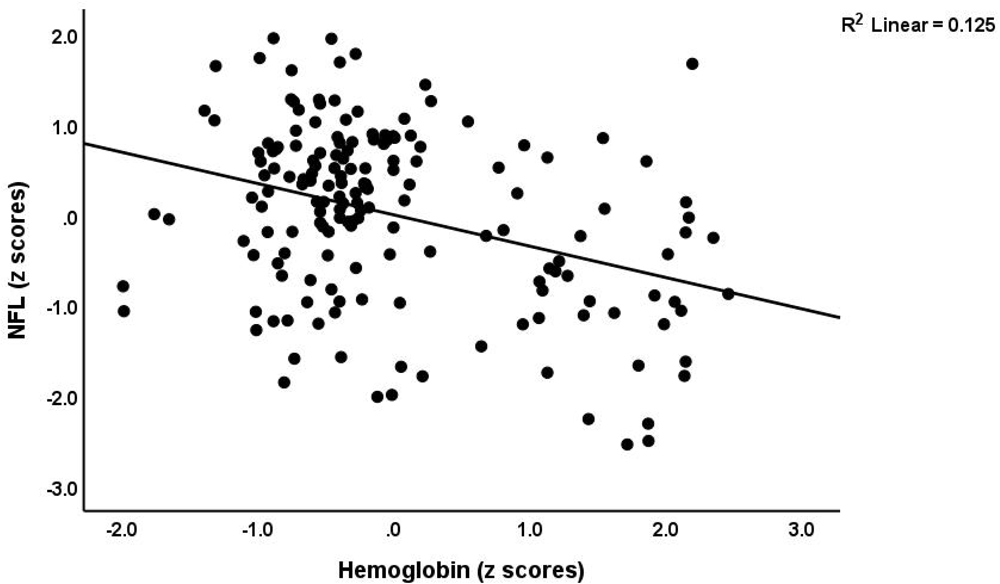
Partial regression of the neurofilament light (NFL) levels on hemoglobin levels, after adjusting for age and sex (p<0.001)

**Figure 4.**
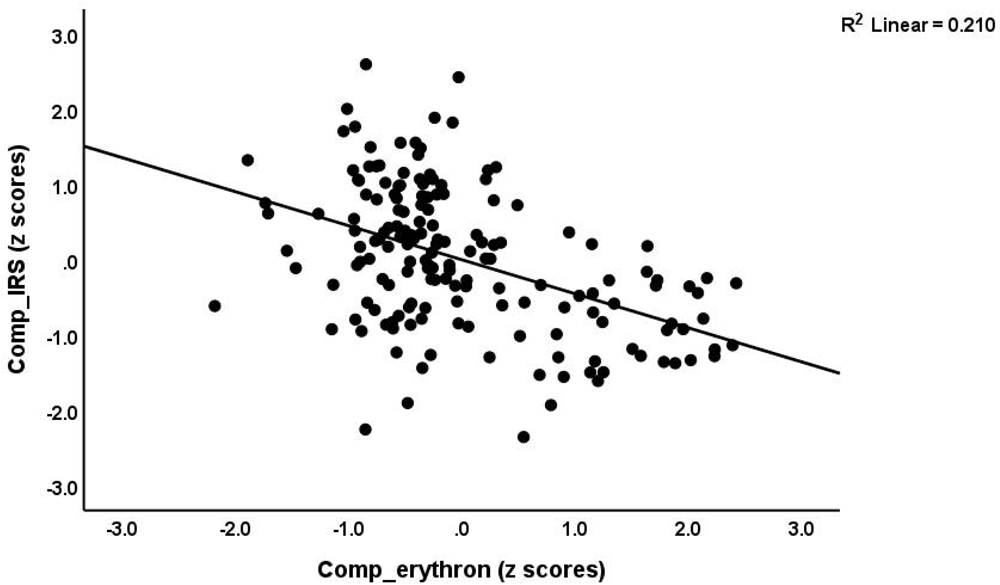
Partial regression of the immune-inflammatory response system (Comp_IRS) index on an erythron index based on hematocrit and hemoglobin measurements (Comp_erythron), after adjustment for age and sex (p<0.001)

## Discussion

### IO, IRS activation, and affective and CFS symptoms due to TDT

The first major finding of this research is that children with TDT have higher total FF, CDI, and SCAS scores in comparison to healthy children. Additionally, there is a significant correlation between the integrated index of severity of affective and CFS symptoms and IRS activation (increased CRP, IL-6 and IL-10, and lowered zinc), IO (increased iron and ferritin), and erythron disorders (lowered hematocrit and hemoglobin).

IRS activation in TDT was identified in the present study by elevated levels of CRP, IL-6, and IL-10 in the serum, in comparison to the control group. Previously, a wide spectrum of immune defects has been observed in patients with affective disorders and CFS (Maes and Carvalho, 2018, Maes et al., 2022a) and β-thalassemic patients (Bazi et al., 2018). Serum concentrations of IL-6, IL-10, and ferritin were all elevated in TDT patients (Politou et al., 2020). TDT has been associated with altered cytokine profiles, modifications in innate immune functions, and the manifestation of a low-grade systemic inflammatory state (Elsayh et al., 2016). Based on existing literature data, immunological abnormalities observed in thalassemia patients may be induced by both the condition and its therapeutic interventions. IO, functional abnormalities of immune system cells caused by chronic inflammation, oxidative stress, multiple blood transfusions, iron chelation therapy, and splenectomy are the most significant contributors to these changes (Gluba-Brzózka et al., 2021).

Depressive and CFS symptoms may result from peripheral activation of IRS pathways influencing the brain via humoral and neural pathways, thereby altering the functions of neural cells, structural and functional connectivity, and ultimately leading to neuropsychiatric symptoms (Kubera et al., 2011, Harrison et al., 2016, Maes et al., 2022a). Neurotoxicity can be caused by various neuro-immune (M1, Th1, and Th17) and neuro-oxidative pathways. This can disrupt crucial neuronal processes, including neurogenesis, neuroplasticity, axonogenesis, dendritic sprouting, and receptor expression (Maes et al., 2022b, Maes et al., 2009). Furthermore, IRS activation disrupts interoceptive signaling and the functional connectivity within the brain networks, which may have ramifications for the etiology of MDD and CFS (Aruldass et al., 2021, Maes et al., 2022b). In addition, heightened concentrations of proinflammatory cytokines trigger an acute phase response in the liver, resulting in augmented CRP production (Sluzewska et al., 1996). This compound further impairs endothelial cells, increasing their permeability and exacerbating the progression of neurodegenerative and cerebrovascular disorders (Aruldass et al., 2021, Hsuchou et al., 2012, Belin et al., 2020, Ge et al., 2013).

Another hypothesis that connects TDT with the development of affective and CFS symptoms is that iron accumulation in certain brain regions has neurotoxic effects. Iron accumulation not only triggers oxidative stress but also exerts an impact on the turnover of neurotransmitters and the expression of neuroreceptors within the brain, consequently influencing behavioral responses (Al-Hakeim et al., 2020, Cutler, 1994, Kim and Wessling-Resnick, 2014). Impaired dopamine metabolism, including dopamine transporters and increased dopamine D2 receptors (Chang et al., 2014), as well as decreased brain serotonin and dopamine concentrations (Elseweidy and Abd El-Baky, 2008), may contribute to the emotional symptoms and behavioral deficits associated with IO. Iron has a direct impact on GABAergic pathways, as do reactive oxygen species catalyzed by iron (Ye et al., 2019), (Accardi, 2016). Given that an overabundance of iron induces the generation of reactive oxygen species (Farina et al., 2013), and that increased oxidative stress is associated with dysfunction in the GABAergic system (Han et al., 2016), the hypothesis proposes that iron accumulation in the brain disrupts GABA homeostasis and impacts affective behavior (Ye et al., 2019). Additionally, fear of separation from family, limited social interaction and isolation, physical and facial deformities, restricted life opportunities, fear of death, maladaptive coping mechanisms, and restrictions in school and outdoor pursuits are all contributing factors to affective disorders in children with TDT (Shah et al., 2020, Koutelekos and Haliasos, 2013).

### Neuronal and astroglial projection toxicity in TDT

The second major finding of the present study is that serum levels of various neuronal and astroglial damage biomarkers are elevated in TDT patients relative to the control group and that these CNS injury biomarkers are positively associated with the severity of affective and CFS symptoms. NFL, GFAP, and NSE were, in descending order of significance, the most pertinent CNS biomarkers. Moreover, there exists a substantial correlation between these markers of CNS injury and IRS biomarkers, as well as copper, IO, or erythron biomarkers.

Elevated NFL levels in patients with neuroinflammatory diseases are indicative of ongoing and acute disease processes as opposed to progressive brain injury (Cantó et al., 2019, Srpova et al., 2021). Moreover, synaptic degeneration may be indicated by an increase in NFL (Zerr et al., 2018), which is a biomarker of injury to cyto-axonal cell structures (Gaetani et al., 2019) in neurodegenerative disorders. The correlation between serum NFL and cognitive performance in healthy individuals suggests that NFL has the capability to detect subclinical microstructural alterations (Beste et al., 2019). Reactive astrogliosis and CNS trauma induce an increase in GFAP expression, which may be translocated into the peripheral circulation via blood-brain-barrier disruption (Yang and Wang, 2015, Hol and Pekny, 2015). Elevated serum GFAP is, therefore, a biomarker for neurotoxicity, specifically injury to neuronal and astroglial cells (Yang and Wang, 2015). Variations in GFAP levels can manifest in response to traumatic injuries, neurodegenerative or neuroinflammatory processes (Bagheri et al., 2015, Abdelhak et al., 2019), neuropathology (Wang and Parpura, 2016), disruption of the blood-brain barrier (Torres-Platas et al., 2016), and brain vascular inflammation. Serum and cerebrospinal fluid NSE elevation is an effective biomarker for neuronal loss and impairment, brain injury, and neuroinflammation (Park et al., 2019, Bagheri et al., 2022, Haque et al., 2018). It is noteworthy that an increase in the release of neuronal biomarkers into the bloodstream (Arslan and Zetterberg, 2023, Negi et al., 2023) through the glymphatic system may result from apoptosis of brain cells (Hablitz and Nedergaard, 2021, Plog et al., 2015).

Patients diagnosed with MDD and those experiencing affective and CFS symptoms because of end stage renal disease exhibit elevated levels of neuronal and astroglial projection markers (Al-Hakeim et al., 2023b, Al-Hakeim et al., 2023a). Cognitive function was found to be inversely correlated with increased NFL in patients with MDD (Bavato et al., 2021). This finding suggests that active neuropathological processes, specifically axonal and synaptic lesions, may be present in these patients (Douillard-Guilloux et al., 2013, Dohm et al., 2017). CSF GFAP levels are elevated in patients with MDD (Michel et al., 2021), and serum GFAP levels may increase as the severity of MDD worsens (Steinacker et al., 2021). Additionally, individuals with neuropsychiatric disorders who do not exhibit any brain radiological abnormalities may have elevated plasma GFAP levels (Esnafoglu et al., 2017). Serum GFAP has been proposed as a potential marker for monitoring astroglial pathology in MDD (Hol and Pekny, 2015, Al Shweiki et al., 2017, Steinacker et al., 2021). It was believed that NSE and GFAP serve as prognostic indicators for high-risk suicide attempters (Bagheri et al., 2022). Furthermore, elevated CRP levels were found to be strongly correlated with increases in these neuronal and astroglial projection injury biomarkers in MDD (Al-Hakeim et al., 2023a). In summary, the findings indicate that the heightened neurotoxicity observed in affective and CFS symptoms might be the result of IRS activation and neuroimmune mechanisms. In the following paragraph, however, we explore additional potential pathways that connect TDT with CNS injury.

An overabundance of iron within the brain has been associated with the onset and progression of neurodegenerative diseases (Bartzokis et al., 2000, Rouault and Cooperman, 2006). Elevated concentrations of iron stimulate the production of reactive oxygen species, which subsequently cause harm to cells and tissues (Nandar et al., 2013, Tuomainen et al., 2007) as well as neurodegenerative disorders (Honda et al., 2004, Xian-hui et al., 2015). Iron toxicity can induce neurodegeneration, which in turn can trigger ferroptosis, a non-apoptotic cell death specific to iron (Kenkhuis, 2022, Zhang et al., 2020). In addition to heightened neuro-oxidative pathways, thalassemia disease may also induce neurodegeneration via mitochondrial dysfunctions (Shaki et al., 2017). Lowered calcium levels, in addition to peripheral inflammation, have the potential to induce neuro-glial projection toxicity (Al-Hakeim et al., 2023a).

### Limitations of the study

The fact that this is a case-control study is an initial limitation; consequently, any potential causal relationships established here must be checked in prospective studies. Furthermore, to make the study even more interesting, we should have assessed a comprehensive range of cytokines and biomarkers associated with neuronal damage in TDT children in conjunction with neuropsychiatric ratings.

## Conclusions

The current investigation showed heightened neurotoxicity among TDT patients, in which immune-inflammatory biomarkers and serum biomarkers of injury to neuronal and astroglial projections are correlated with the intensity of affective and CFS symptoms. IO may induce both immune activation and increased neurotoxicity in TDT. Activated IRS pathways, increased neurotoxicity, and subsequent injury to neuronal and astroglial projections are, therefore, new drug targets for the treatment and prevention of affective and CFS symptoms caused by IO and TDT, according to this study. In addition to enhancing neuroprotection, novel pharmaceutical interventions should be developed to target neurotoxicity and IRS activation.

## Data Availability

All data produced in the present study are available upon reasonable request to the authors

## Acknowledgment

We thank the staff of the Thalassemia Unit at Al-Zahra’a Teaching Hospital-Najaf city-Iraq for their help in the collection of samples. We also acknowledge the work of the highly skilled staff of the Asia Laboratory in measuring the biomarkers.

## Author’s contributions

All the contributing authors have participated in the preparation of the manuscript.

